# Edge Time Series Components of Functional Connectivity and Cognitive Function in Alzheimer’s Disease

**DOI:** 10.1101/2023.05.13.23289936

**Authors:** Evgeny J. Chumin, Sarah A. Cutts, Shannon L. Risacher, Liana G. Apostolova, Martin R. Farlow, Brenna C. McDonald, Yu-Chien Wu, Richard Betzel, Andrew J. Saykin, Olaf Sporns

## Abstract

Understanding the interrelationships of brain function as measured by resting-state magnetic resonance imaging and neuropsychological/behavioral measures in Alzheimer’s disease is key for advancement of neuroimaging analysis methods in clinical research. The edge time-series framework recently developed in the field of network neuroscience, in combination with other network science methods, allows for investigations of brain-behavior relationships that are not possible with conventional functional connectivity methods. Data from the Indiana Alzheimer’s Disease Research Center sample (53 cognitively normal control, 47 subjective cognitive decline, 32 mild cognitive impairment, and 20 Alzheimer’s disease participants) were used to investigate relationships between functional connectivity components, each derived from a subset of time points based on co-fluctuation of regional signals, and measures of domain-specific neuropsychological functions. Multiple relationships were identified with the component approach that were not found with conventional functional connectivity. These involved attentional, limbic, frontoparietal, and default mode systems and their interactions, which were shown to couple with cognitive, executive, language, and attention neuropsychological domains. Additionally, overlapping results were obtained with two different statistical strategies (network contingency correlation analysis and network-based statistics correlation). Results demonstrate that connectivity components derived from edge time-series based on co-fluctuation reveal disease-relevant relationships not observed with conventional static functional connectivity.

## Introduction

Various neuroimaging modalities can now capture multiple facets of Alzheimer’s disease (AD), offering tools for characterization and understanding of disease impact on brain and cognitive functions. Such understanding is important as over 6 million people are affected by AD in United States alone, a number that is projected to rise to over 12 million by 2050 according to the 2022 Alzheimer’s Association Annual Report. Pathological hallmarks of AD, beta-amyloid plaques and hyperphosphorylated tau tangles, have been imaged *in vivo* with positron emission tomography, showing increasing accumulation of both as disease severity progresses (Therriault et al. 2022). Accumulation of tau and its spread have also been shown to occur within the functional network organization of the brain (Franzmeier et al. 2020; Franzmeier et al. 2019). These networks were identified in early functional magnetic resonance imaging (fMRI) studies of task-based and resting-state connectivity (Buckner et al. 2008; Fox et al. 2006; Yeo et al. 2011). Since then, numerous fMRI studies in AD have reported alterations in the properties of resting state networks (RSNs) such as their strength (Dai et al. 2019; Dai et al. 2015) and interconnectivity (Forouzannezhad et al. 2019). Changes in these network properties have also been related to cognitive function (Chumin et al. 2021; Contreras et al. 2019) and existing biomarkers (Smith et al. 2021; Veitch et al. 2019).

In recent years, in parallel with the improvement in the temporal resolution of fMRI, studies of the temporal dynamics of the brain and its RSNs have emerged (Hutchison et al. 2013; Lurie et al. 2020). Even over the relatively short duration of a typical fMRI scan, brain functional networks exhibit significant dynamic fluctuations, and this observation raises the question whether time points differentially contribute/relate to neuropsychological outcomes of interest. This is supported by literature investigating brain states, where clustering algorithms were used to group functional connectivity (FC) patterns of activity (Calhoun et al. 2014; Cohen 2018). AD-related alterations in the dynamics of FC are marked by reduced internetwork connectivity (Schumacher et al. 2019), which is related to cognitive function (Franzmeier et al. 2017). Additionally, the emergence and duration of these states, as well as the transition between them, has been shown to be different in AD relative to other diagnostic groups (Schumacher et al. 2019). Such methods divide the data into non- or partially overlapping temporally continuous subsets (windows) to study FC-cognition relationships in AD. However, over the duration of a resting-state scan, it is likely that individual time points differentially relate to neurocognitive outcomes and behaviors. In this case, methods that assess functional properties at single repetition time (TR, a single fMRI time point) resolution are better suited to probe these relationships. To date, no methods have been employed in clinical AD that probe brain-behavior at single-TR resolution.

A method to probe single-TR connectivity dynamics has recently been proposed by Faskowitz et al. (2020), which relies on ‘temporal unwrapping’ of the Pearson correlation conventionally used to estimate FC, to yield moment-to-moment co-fluctuations. Computed as the elementwise product of regional blood-oxygen-level-dependent (BOLD) signals, co-fluctuations are represented as an edge (connection) by time matrix of edge time-series (ETS). This approach offers an intuitive interpretation of the ongoing dynamics in the brain and has been employed to probe modular/community structure (Faskowitz et al. 2020; Jo et al. 2021), individual variability (Betzel et al. 2022; Cutts et al. 2022; Sasse et al. 2022), and disease-related alterations in brain function (Idesis et al. 2022; Zamani Esfahlani et al. 2022). Previous work on ETS in young healthy individuals from the Human Connectome Project (Van Essen et al. 2013) dataset has shown that FC can be approximated from a subset of scan time points with highest co-fluctuations (Zamani Esfahlani et al. 2020) and that identifiability of individuals was improved by focusing on subsets of time points of intermediate co-fluctuation magnitude (Cutts et al. 2022). We have previously shown a relationship between a time-varying measure of RSNs connectivity and cognitive function (using a sliding window approach) in a cross-sectional sample spanning the AD diagnostic continuum (Chumin et al. 2021). Here, we hypothesized that the application of ETS to group temporally dispersed time points into FC components (FCc), thus separating time points based on co-fluctuation magnitude, would reveal relationships between connectivity and neuropsychological measures that are not detectable and perhaps obscured in conventional ‘full FC’.

## Methods

### Indiana Alzheimer’s Disease Research Center (IADRC) Sample

Data were collected at the IADRC, as part of the Indiana Memory and Aging Study, at the Indiana University School of Medicine. Valid datasets (as determined by quality control of image preprocessing) for 152 individuals were included in the present study (Table 1). The sample consisted of 53 cognitively normal controls (CON; no cognitive concerns), 47 subjective cognitive decline participants (SCD; significant cognitive concerns despite normative test performance), 32 mild cognitive impairment participants (MCI; cognitive performance below the normal range), and 20 Alzheimer’s disease patients (ALZ). Demographics and neuropsychological domain group comparisons were carried out with a one-way analysis of variance with Tukey-Kramer post hoc tests or chi-squared tests, as appropriate. Informed consent was obtained from all participants or their representatives, and all procedures were approved by the Indiana University Institutional Review board in accordance with the Belmont report. A portion of participant data from this sample have been used in previous publications (Chumin et al. 2021; Contreras et al. 2019).

**Table 1.**
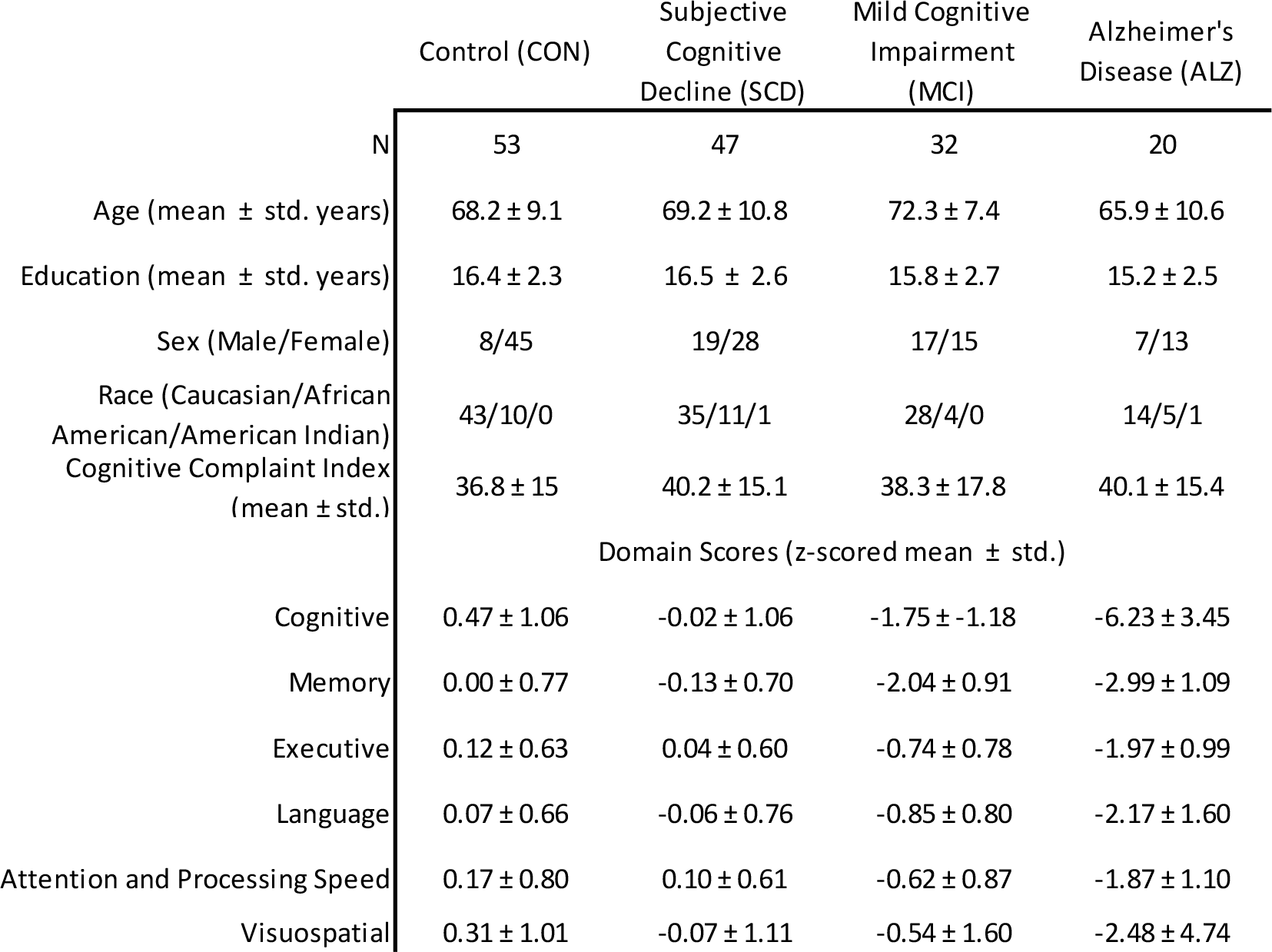
Demographic and Neuropsychological Characteristics. Data are shown as counts or mean and standard deviation (std.). Age, years of education, and race distribution did not significantly differ among groups. There was a significant difference in distributions of sex (Χ^2^(3, N=152) = 14.6, *p* < 0.01). All six domains showed a significant effect of group (ANOVA, *p* < 0.0001). The four numbers in the right column next to domain names correspond to number of missing data points for each group. Domain scores were derived from the following: Cognitive – Montreal Cognitive Assessment (total score); Memory – Logical Memory (immediate and delayed), CERAD Word List Learning (immediate and delayed), Selective Reminding Test (delayed), 7/24 Spatial Recall Test (immediate & delayed), Rey Auditory Verbal Learning Test (RAVLT; immediate and delayed), Craft stories (immediate and delayed), and Benson Complex Figure (delayed recall); Executive – Digit Span (backwards), Trail Making B, Digit Symbol Substitution, Wisconsin Card Sorting Test (categories & perseverations), Controlled Oral Word Association (COWA), Stroop (Word, Color, and Color-Word scores), UDS3 Letter Fluency; Language - Animal Fluency, Vegetable Fluency, Boston Naming Test, IU Token Test, COWA, Multilingual Naming Test, UDS3 Letter Fluency; Attention and Processing Speed - Digit Span (forward & backward), Trail Making A and B, Digit Symbol, Stroop (Word, Color, & Word/Color); Visuospatial - Benson Complex Figure (copy), Judgement of Line Orientation, Block Design.

### IADRC Neuropsychological Scores

Participants completed neuropsychological testing as part of the Uniform Dataset 3.0 (Weintraub et al. 2018), as well as site-specific additional tests. Six domain composite scores were calculated from the following: (1) Cognitive – Montreal Cognitive Assessment (total score) (Nasreddine et al. 2005), (2) Memory – Logical Memory (immediate and delayed) (Wechsler 1987), CERAD Word List Learning (immediate and delayed) (Petersen et al. 1992), Selective Reminding Test (delayed), 7/24 Spatial Recall Test (immediate & delayed), Rey Auditory Verbal Learning Test (RAVLT; immediate and delayed) (Schmidt 1996), Craft stories (immediate and delayed) (Craft et al. 1996), and Benson Complex Figure (delayed recall) (Possin et al. 2011), (3) Executive – Digit Span (backwards) (Ivnik et al. 1992), Trail Making B (Steinberg et al. 2005), Digit Symbol Substitution, Wisconsin Card Sorting Test (categories & perseverations), Controlled Oral Word Association (COWA), Stroop (Word, Color, and Color-Word scores), UDS3 Letter Fluency (Weintraub et al. 2018), (4) Language - Animal Fluency, Vegetable Fluency, Boston Naming Test, IU Token Test, COWA, Multilingual Naming Test, UDS3 Letter Fluency, (5) Attention and Processing Speed - Digit Span (forward & backward), Trail Making A and B, Digit Symbol, Stroop (Word, Color, & Word/Color), (6) Visuospatial - Benson Complex Figure (copy), Judgement of Line Orientation, Block Design. To generate the composite scores, all scores were first adjusted for age, sex, and years of education, *z-*scored relative to a sample of independent (non-overlapping) cognitively normal controls, and then the z-scores were averaged within each domain as described previously (Chumin et al. 2021; Contreras et al. 2019).

### IADRC Image Acquisition and Processing

Both image acquisition and preprocessing have been described in detail previously (Chumin et al. 2021). Participants were scanned on a Siemens 3T Prisma Scanner (Siemens, Erlangen, Germany) with a 64-channel head coil. A T1-weighted, whole-brain magnetization prepared rapid gradient echo (MPRAGE) volume was acquired with parameters optimized for the Alzheimer’s Disease Neuroimaging Initiative (ADNI 1 & 2; http://adni.loni.usc.edu): 220 sagittal slices, GRAPPA acceleration factor of 2, voxel size 1.1×1.1×1.2 mm^3^, duration 5:12 minutes. Two spin-echo echo-planar imaging (12 sec each, TR = 1.56 sec, TE = 49.8 ms, flip angle 90°) volumes were acquired with reverse phase encoding directions for distortion correction. Resting-state functional MRI (rs-fMRI) data were acquired with a gradient-echo echo-planar imaging sequence with a multi-band factor of 3, 10:07 min scan time, and TR of 1.2 sec, resulting in 500 time points. Other relevant parameters were TE = 29 ms, flip angle 65°, 2.5×2.5×2.5 mm^3^ voxel size, and 54 interleaved axial slices. During the scan, participants were instructed to remain still with eyes closed and to think of “nothing in particular.”

Data were processed with a pipeline developed in-house, implemented in Matlab (MathWorks, version 2019a; Natick, MA), utilizing the Oxford Centre for Functional MRI of the Brain (FMRIB) Software Library (FSL version 6.0.1) (Jenkinson et al. 2012), Analysis of Functional NeuroImages (AFNI; afni.nimh.nih.gov), and ANTS (http://stnava.github.io/ANTs/) packages. This pipeline was developed and optimized for the Siemens scanner data acquired at Indiana University School of Medicine following recommendations in Lindquist et al. (2019); Parkes et al. (2018); Satterthwaite et al. (2013).

All processing was carried out in each participant’s native space. T1 volumes were denoised (Coupé et al. 2008), bias field corrected (FSL), and skull stripped (ANTS). rs-fMRI data were first distortion corrected (FSL *topup*), motion corrected (*mcflirt*), and normalized to mode 1000. Nuisance regressors were removed from the data with use of ICA-AROMA (Pruim et al. 2015), aCompCor (Muschelli et al. 2014), and global signal regression. Data were then demeaned, detrended, and bandpass filtered (0.009–0.08 Hz). Finally, 30 time points were removed from the beginning and end of the scan to remove edge artifacts introduced by bandpass filtering and ensure equal binning (see below). Relative frame displacement output by *mcflirt* was used as an index of in-scanner motion.

### Cortical Parcellation and Time-Series Extraction

A 200-cortical region brain parcellation was spatially aligned with each subjects’ rs-fMRI via the following: (1) linear (6 and 12 degrees of freedom; FSL *flirt*) and nonlinear (FSL *fnirt*) registration of T1 volume, with inverse transformation applied to the Schaefer et al. (2018) 200 node (cortical region) parcellation, (2) dilation and application of a gray matter mask, and (3) registration of T1 to the mean rs-fMRI volume with a linear white matter boundary-based registration (FSL *flirt bbr* cost function). Nodal time-series were then extracted as the mean time course across voxels in each region. This was repeated for the 300-node Schafer parcellation for supplementary assessment of the impact of parcellation scale.

### Edge Time-Series and FC Components

ETS were computed as the frame-by-frame product of the z-scored BOLD time-series for all region pairs (19,900 unique edges) (Faskowitz et al. 2020; Jo et al. 2021), resulting in an edge by time matrix of moment-to-moment co-fluctuations (Figure 1A-B), which is analogous to temporally unwrapping the Pearson correlation (the mean over time of ETS is equal to the conventional “full FC” or “static FC”). The ETS matrix was then used to compute root-sum-square (RSS) at each time point as an index of global co-fluctuation amplitude (Figure 1C).

**Figure 1.**
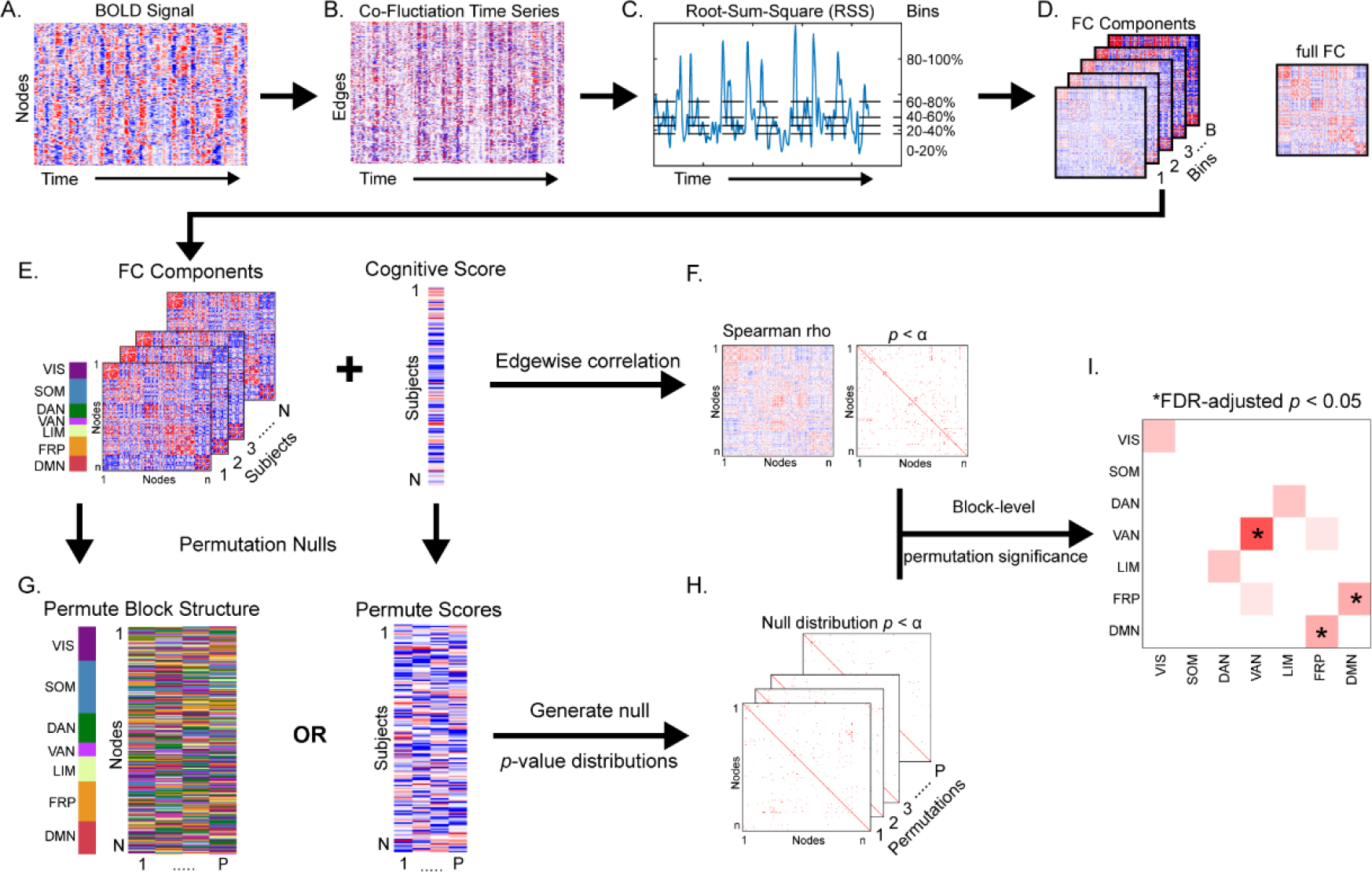
Edge time-series, functional connectivity (FC) components, and Network Contingency Correlation. (A) Blood-oxygen-level-dependent (BOLD) time-series are z-scored and multiplied for all node pairs to yield (B) edge time-series that describe moment-to-moment co-fluctuation among regions. (C) Root-sum-square (RSS), an index of total co-fluctuation magnitude, is computed at each time point and used to rank and parse time points into equally sized bins, with the mean within each bin corresponding to (D) an FC component. Additionally, full FC is computed as the mean of all time points and is equivalent to Pearson correlation. FCc estimates are correlated at each edge with cognitive domain scores of interest to generate (F) a correlation and a *p*-value matrix. (G) A permutation null (either scrambling the network block structure or cognitive domain scores) is then employed to generate (H) a set of null matrices. (I) Network-block level significance is then computed as a permutation *p-*value (one minus the number of times the count of significant edges within a block in empirical data exceeded null data). *p*-value is then adjusted for number of blocks tested with false discovery rate (FDR) correction. VIS – visual, SOM – somatomotor, DAN – dorsal attention, VAN – ventral attention, LIM – limbic, FRP – frontoparietal, DMN – default mode network.

RSS ranked time points were then divided into 5 equally sized bins (43 TRs per bin, ∼52 second of noncontiguous data). A 10-bin set was also generated to assess the influence of number of bins. The mean edgewise co-fluctuation within each bin was then computed and is referred to as an FC component (FCc; Figure 1D). Each component is thought of as a representation of co-fluctuation within its RSS band, and we hypothesized that different FC components would differentially associate with neurocognitive domains.

### Network Contingency Correlation (NCC) Analysis Framework

To identify relationships between neuropsychological domains and FC (full and RSS components), a modified network contingency analysis (NCA) (Contreras et al. 2019; Sripada et al. 2014) was employed. The NCA framework uses a *t*-test to compute edge-level group differences, then counts the number of significant edges within blocks (i.e., RSNs) and determines block-level significance relative to a permuted data null. This method sidesteps the limitation of mass univariate testing, without averaging data and diluting potential effects. Here, in formulating NCC, the group inference via *t*-test was replaced by a Spearman correlation, quantifying the relationship between individual network edges and behavioral measures. The NCC procedure is applied as follows: (1) edgewise correlations are computed between FC (full or component) and a behavioral domain score, which yields a matrix of correlation coefficients and a binary significance matrix (here the edge-level threshold was set at *p* < 0.01; Figure 1E-F), (2) data are permuted (we tested two null models: a block permutation where RSNs assignment are scrambled and a score permutation where the behavioral scores are scrambled across subjects; Figure 1G) and a distribution of null significance matrices is generated (5,000 permutations, Figure 1H)), (3) the block structure is imposed on the empirical and null significance networks (here we used the 7 canonical RSNs described Yeo et al. (2011) (visual, somatomotor, dorsal and ventral attention, limbic, frontoparietal, and default mode) with node assignments provided in Schaefer et al. (2018)) and the number of significant edges is counted for all within- and between-RSN blocks, and (4) a block-level significance *p*-value is defined as one minus the fraction of instances where the count of significant edges in a block exceeded the null, followed by a false discovery rate (FDR) adjustment for number of blocks (7 within and 21 between RSN) at *q* < 0.05 (Figure 1I). An exploratory run of NCC was also performed without imposing a block structure (i.e., treating all nodes as belonging to one block) and using a network-based statistics largest connected component-based correction with an initial edge-level threshold of *p* < 0.01 (Zalesky et al. 2010).

## Results

### Demographic group comparisons

No differences in age (ANOVA, F(3,148)=2.07, *p* > 0.05), education (F(3,148)=1.54, *p* > 0.05), or race (Χ^2^(3, N=152) = 5.5, *p* > 0.05) were observed. There were proportionally more female participants in the CON and SCD groups (Χ^2^(3, N=152) = 14.6, *p* < 0.01). All neuropsychological domains showed a significant main effect of group (ANOVA: Cognitive F(3,142) = 87.9, Memory F(3,142) = 85.4, Executive F(3,140) = 39.6, Language F(3,140) = 30.1, Attention and Processing Speed F(3,141) = 28.7, and Visuospatial F(3,139) = 8.7, all *p* < 0.0001). Post hoc testing showed that for 4 domains (not including memory and visuospatial) only CON v. SCD comparisons were not significant (*p* > 0.05). For the memory domain CON v. SCD and MCI v. ALZ were not significant (*p* > 0.05). Finally, only the pairwise comparisons against the ALZ group were significant for the visuospatial domain (*p* < 0.05).

### Characterization of RSS quantile FCc

Group-averaged FC and FCc matrices are shown in Figure 2. FC (correlation matrices; Figure 2A) are bounded [-1 1], while FCc matrices of average co-fluctuation within RSS quantiles are not, as evident by increasing amplitude with increasing RSS quantile. To determine if there is unique information within each FCc, we cross-correlated all participants to assess their similarity (Figure 3A). Full FC showed highest correlation values approaching Pearson r values of +0.6. Subject cross-correlation qualitatively increased for increasing RSS quantile FCc; however, they stayed below full FC, suggesting greater relative inter-subject variability. No relationship to in-scanner motion (frame displacement) was found for single time point RSS values in this sample (Pearson r = 0.05, Figure 3B). Comparisons of group average FCc (visualized in Figure 2B-F) showed that ALZ group FCc were least correlated with the other 3 diagnostic groups and that within group, FCc from distant RSS quantiles had lower correlation values (Figure 3C). Finally, as shown in previous work where top 5% RSS time points were highly correlated with FC (Zamani Esfahlani et al. 2020), when correlating quantile FCc to full FC, components derived from greater RSS percentiles (higher co-fluctuation time points) were more similar to full FC (Figure 3D).

**Figure 2.**
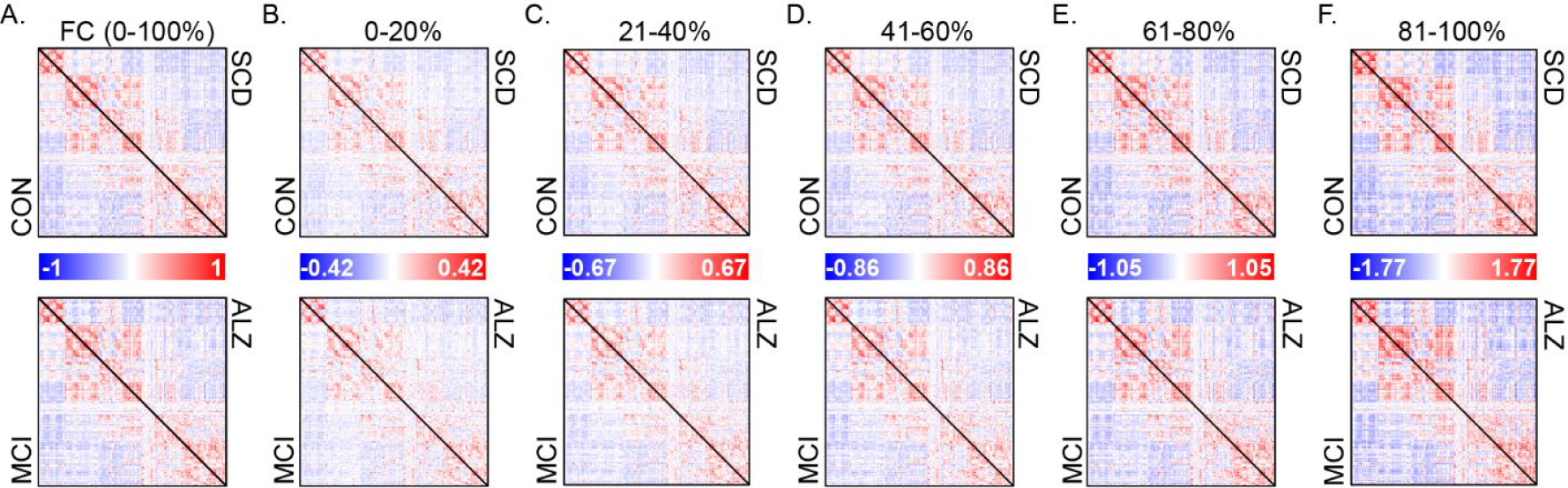
Group averaged full functional connectivity (FC) and its components. (A) Average group full functional connectivity computed as Pearson correlation, which is equivalent to the average co-fluctuation of all time points. Each triangle within a matrix is a group averaged network of unique edges: Controls (CON) – top matrix lower triangle, subjective cognitive decline (SCD) – top matrix upper triangle, mild cognitive impairment (MCI) – bottom matrix lower triangle, and Alzheimer’s disease (ALZ) – bottom matrix upper triangle. (B-F) Group averaged FC components, each comprised of 20% of root sum square (RSS) ranked time points, with 0-20% corresponding to lowest RSS amplitude bin, and 80-100% to the highest amplitude.

**Figure 3.**
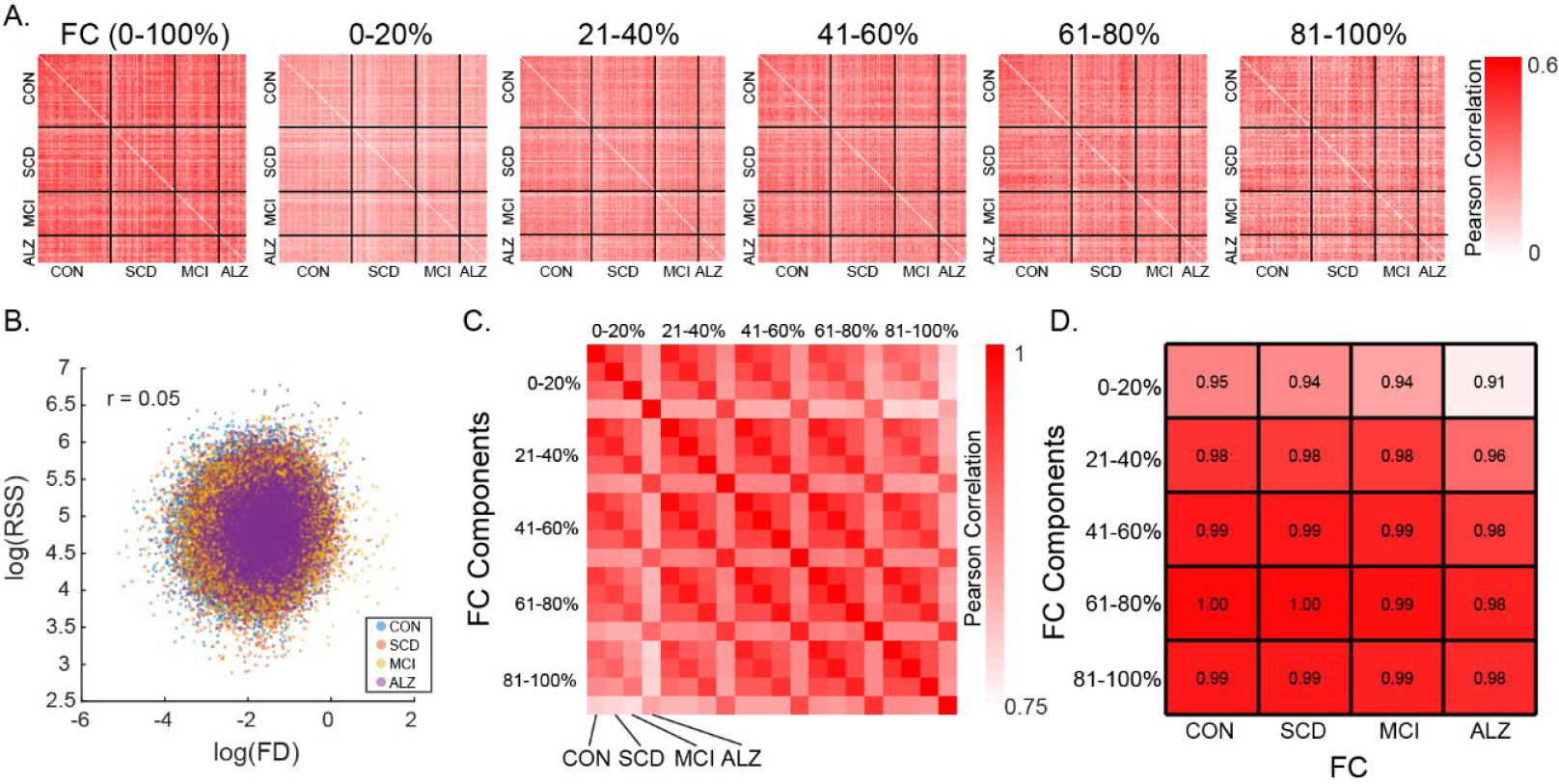
Similarity among FC and its components and relationship of root-sum-square (RSS) with in-scanner motion. (A) Cross-correlation among participants (ordered by diagnostic group) of full FC and the 5 RSS FC components. (B) Scatter correlation (Pearson r) of RSS and frame displacement (FD) shown in log scale and colored by diagnostic group. (C) Cross-correlation (Pearson r) of group averaged FC components ordered by increasing RSS bin and by group within each bin. (D) Correlation of group averaged full FC to each of the RSS FC components split by group. CON – Controls, SCD – subjective cognitive decline, MCI – mild cognitive impairment, ALZ – Alzheimer’s disease.

### Network Contingency Correlation

Analysis of FC/FCc relationships with neuropsychological domain scores showed that permutation of domain scores was a more conservative strategy compared to RSN block label permutation (Supplementary Figures 1, 2). While the purpose of the score permutation null is to destroy the subject connectivity-behavior relationship to test if number of significant edges in a block is meaningful, the purpose of the block structure permutation null is to assess if the distribution of significant edges is clustered within a particular block. Therefore, by focusing only on RSN blocks that were identified in both strategies, we determine whether the number and the distribution of significance in a block is robust. The cognitive domain was the only one to show significant relationships for both FC and FCc bins for the ventral attention network (RSS quantiles 40-60% and 60-80%, Figure 4A, D-E).

**Figure 4.**
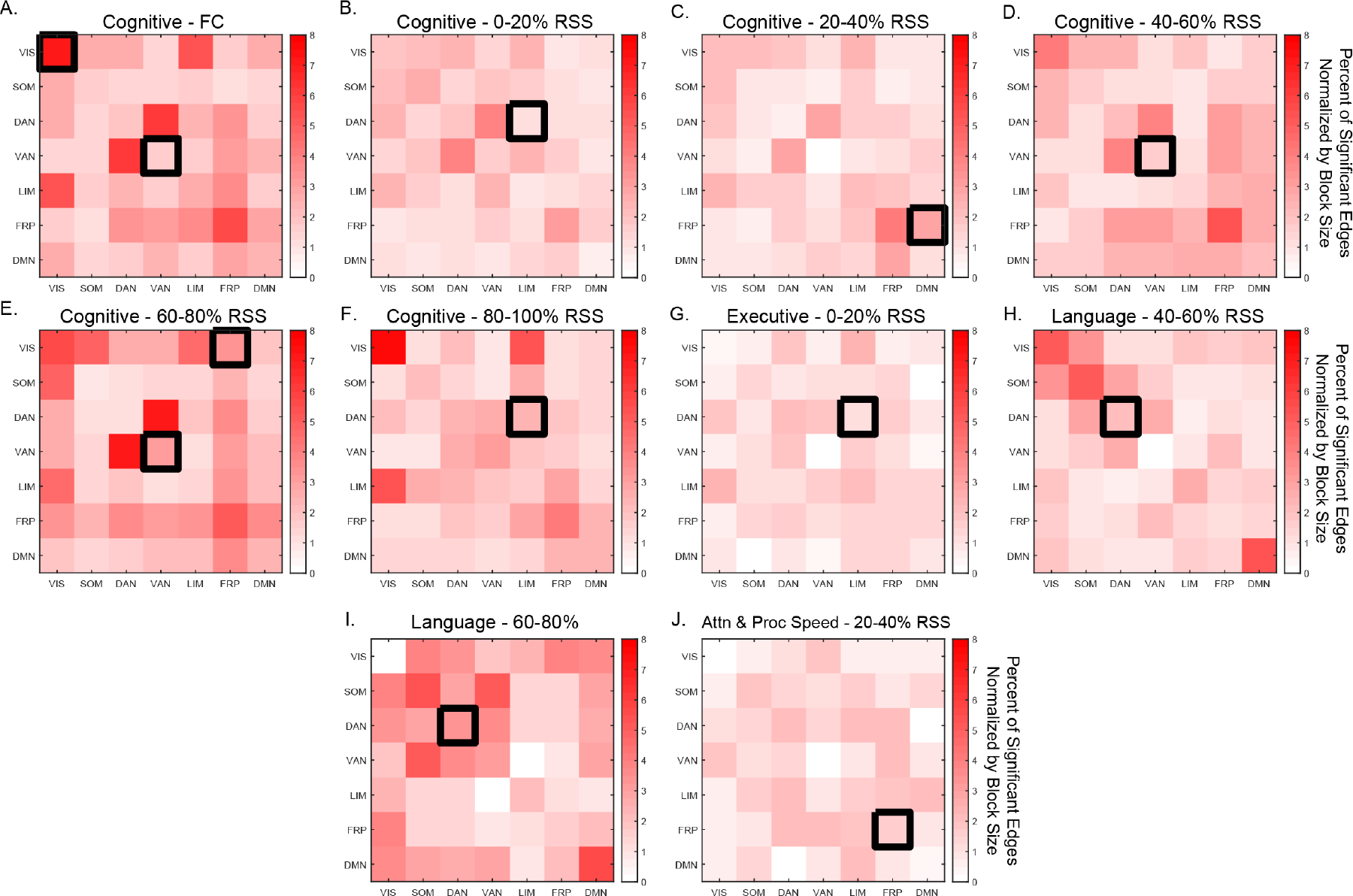
Network Contingency Correlation (NCC) resting state network block-level results. Matrices show percent of edges by block (normalized by size of block) that passed initial uncorrected edge-level significance of *p* < 0.001. Black boxes denote block-level significance of *p*_*FDR*_ < 0.05, with 5,000 NCC permutations. Only blocks that were significant against both permutation nulls are shown. Upper and lower triangular of matrices are identical; significance is only shown on the upper triangle. FC – functional connectivity, RSS – root sum squared. Attn & Proc Speed – attention and processing speed. Resting state networks: VIS – visual, SOM – somatomotor, DAN – dorsal attention, VAN – ventral attention, LIM – limbic, FRP – frontoparietal, DMN – default mode network.

Association between the visual system and the cognitive domain was also observed, but only for FC (Figure 4A). The remaining associations with the cognitive domain were identified in between system interaction blocks: limbic-dorsal attention (0-20% and 80-100% RSS FCc, Figure 4B, F), frontoparietal-default mode interaction (20-40% FCc, Figure 4C), and the frontoparietal-visual interaction blocks (60-80% FCc, Figure 4E). In addition, the executive function domain associated with the limbic-dorsal attention interaction block (0-20% FCc, Figure 4G), the language domain associated with the dorsal attention block (40-60% and 60-80% FCc, Figure 4H-I), and the attention and processing speed domain associated with the frontoparietal system (20-40% FCc, Figure 4J). Across both null strategies and all comparisons, the upper bound of the percent of significant edges (normalized by the size of the RSN block) was 8%. While this is a relatively small fraction of total edges within a network block, each edge passed the initial *p* < 0.01 significance threshold, with the blocks achieving FDR-adjusted significance of *p* < 0.05 (corrected for 7 within and 21 between RSN blocks tested for each domain). To assess the impact of number of bins and number of nodes, the analysis was repeated with 10 FCcs as well as with 5 FCc from 300-node data, with results showing a great degree of similarity (Supplementary Figures 3 and 4). In the 10 FCc binned data, largely the same RSS percentile components and neuropsychological domains were identified. Analysis of 300 node networks also showed overlap, highlighting the same RSN systems (ventral and dorsal attention) and domains (Cognitive, Executive, Language, and Attention and Processing Speed), however, the low RSS bins (0-40%) were not associated with the cognitive domain in the 300 node data.

### Exploratory Network-Based Statistics (NBS) Component Analysis

NBS employs permutation testing to identify connected clusters of nodes above a random null. Six clusters were identified: three associated with the cognitive domain, including full FC (Figure 5A), 40-60% RSS range-derived FCc (Figure 5B), and 60-80% FCc (Figure 5C), one association between the executive domain and 60-80% FCc (Figure 5D), and two associations between the language domain and 40-60% FCc, as well as 60-80% FCc, edges (Figure 5E-F). Degree distributions show that these are extensively interconnected components with multiple neighbors to most nodes, while the matrices show that they are composed of both positive and negative correlations with behavior. These components differ in their composition depending on FC component and domains being compared and are largely characterized by positive within-RSN and negative between-RSN edges (although this is not ubiquitous; see Figure 5C and 5D for notable examples of positive associations between RSN connectivity and cognitive and executive domains, respectively). The same connectivity/neuropsychological domain pairing were also seen in the 300-node networks, with 4 additional clusters: 0-20% FCc with cognitive and executive function domains, FC and the language domain, and 60-80% RSS FCc and the Memory domain (Supplementary Figure 5).

**Figure 5.**
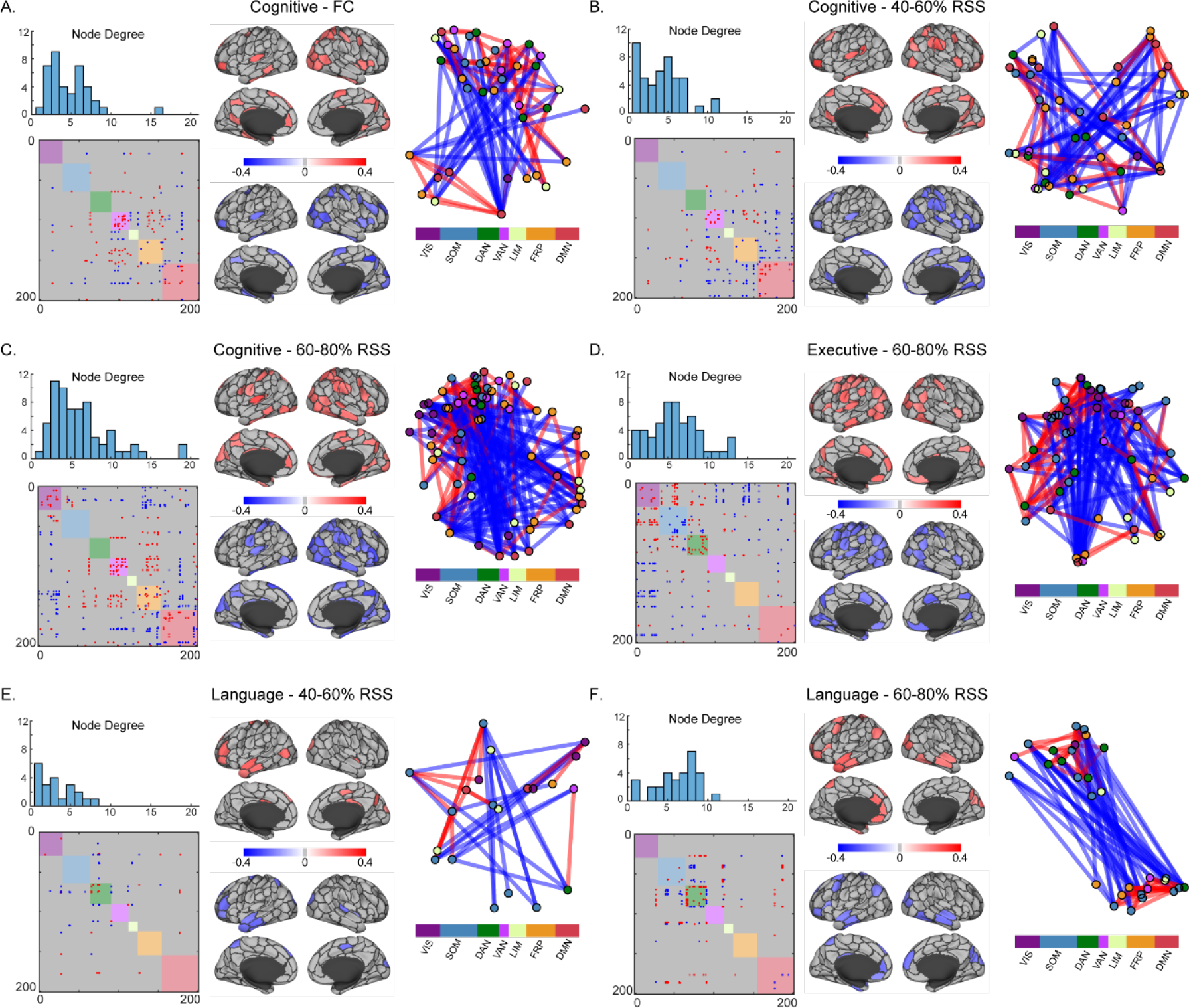
Block-free Network-Based Statistics significant FC component-neuropsychological domain correlations. For each of the six significant components identified across six neuropsychological domains and five FC components and full FC, (upper-left) histograms show nodal degree distributions, (lower-left) matrices show binary positively and negatively correlated edges within each component, (middle) Schaefer 200-node cortical parcellations show average positive and negative edge correlations for component nodes, (right) a force-directed diagram from component nodes colored by Yeo systems and edges colored as positively (red) or negatively (blue) correlated with the neuropsychological domain of interest, shown using an inverse weighting visualization, such that higher edge weights yield shorter distances. System labels are VIS-Visual, SOM-Somatomotor, DAN-Dorsal Attention, VAN-Ventral Attention, LIM-Limbic, FRP-Frontoparietal, DMN-Default Mode. Root-sum-square (RSS) percentiles refer to boundaries of RSS values from which FC components were computed.

## Discussion

The identification of robust and reproducible brain-behavior relationships has significant implications for neuroscience. Their reliable detection in noninvasive fMRI data presents a tremendous challenge, as their expression may be highly time and context dependent. Here, in the context of AD, we presented findings from application of a single time point fMRI framework (edge time-series), to assess whether certain moments in time (as indexed by root sum squared (RSS) ranked co-fluctuations), are preferentially correlated with neuropsychological performance in a cross-sectional sample that spans the AD diagnostic spectrum. Employing block-level inference we identified RSNs and RSN interactions of FC components (FCc) that correlated with neuropsychological function domains. Aside from full FC of the ventral attention network and its correlation with the cognitive function domain, the identified relationships were not significant when using conventional/static FC (cross-correlation of the full time-series). The appearance of the association between cognitive function and FC of the ventral attention system confirms previous findings obtained with sliding-window dynamic FC that its temporal properties are robustly related to cognitive function (Chumin et al. 2021).

Single-TR decomposition of fMRI edge time series allows for many possible strategies to generate FCc. Here we divided the fMRI time series into 5 equal bins based on ranking time points by their overall co-fluctuation amplitude (RSS), yielding 5 FCc. We found that the cognitive domain showed the highest number of associations with FCc RSN blocks across all 5 RSS bins, with notable RSN blocks including the attentional systems (ventral and dorsal attention-limbic interaction). This is consistent with functional interactions of attentional systems and cognitive function, resource recruitment, and reserve (Anthony and Lin 2018; Bastin et al. 2012; Gordon et al. 2015; Zhang et al. 2015). Blocks that included the dorsal attention system were also identified to relate to executive and language domains. Additionally, the frontoparietal control system and its interactions with other RSNs appears for cognitive and attention domains at two RSS bins. These relationships are not found in the full FC matrix, ‘obscured’ by the high co-fluctuation time points that drive the RSN block structure observed in human rs-fMRI.

There is a distinction between the broadly applied sliding window methods in AD and the ETS approach (Faskowitz et al. 2022; Lurie et al. 2020). Both operate on subsets of data; however, ETS rely on a single time point decomposition of the Pearson correlation, making no assumptions about the duration of underlying dynamics. Operating on a shorter time scale with ETS is beneficial, as it better characterizes the ongoing dynamics and has a narrow autocorrelation structure. A second distinction between ETS and previous methodology is that here connectivity of specific systems is being investigated. To date, studies that have employed sliding window FC to study AD have relied on clustering of group connectivity patterns into states. This is a data reduction strategy, as properties of connectivity states (i.e., frequency and dwell time) are then used as predictors/outcomes for statistical analysis. While this is a reasonable approach, it tests whether a property of a state, which is a whole-brain descriptor, is related to behavior of interest. Systems within the brain are likely to differentially relate to behavior, so properties of states, which are whole network descriptors, are unlikely to robustly relate to behaviors. Therefore, a focus on subsystems within the brain, which share some intrinsic properties, may allow us to identify robust behavioral correlates.

The strategy undertaken here (system/block level inference) is also one of data reduction, aimed at avoiding mass univariate tests to obtain interpretable outcomes. However, unlike sliding window, data reduction is caried out during inference through application of frequency statistics. The network contingency analysis proposed by Sripada et al. (2014) was developed to identify block-level group differences between networks, by testing whether the number of significantly different edges in a block exceeded the number expected to occur by chance (with permutation testing).

For such a method to be applicable/interpretable, the a priori defined blocks must share intrinsic properties of interest, with the 7 cortical RSNs fitting that definition. We chose not to include subcortical regions in the above analyses due to the heterogeneous functional nature of its individual regions, as treating whole subcortex as a single block would not be appropriate/interpretable. Therefore, for testing associations with only the 7 cortical systems, we modified the NCC framework for use with correlations, proposing two permutation null strategies for assessment of block-level brain-behavior relationships. The two strategies each permute one of two variables of interest (either FC or behavior), testing whether the observed edge relationships preferentially cluster within a block or if the number of edge-level correlation exceeds the null distribution, respectively. The joined significance against the two models then describes whether the relationships within an RSN block are significant both in number and spatial distribution within the network.

A similar approach to linking fMRI to behavioral and/or neuropsychological measures as the one employed here is connectome-based predictive modeling (CPM), which relies on a cross-validation strategy to build behavior predictive models, by first selecting a subset of edges with the strongest relationship to the behavior of interest (Finn et al. 2015; Shen et al. 2017). Yet it is distinct from the methodology herein as it requires training and testing sets for cross validation. Identified edges can then be qualitatively described in terms of which regions/systems they are comprised of. This strategy has been applied in AD (Lin et al. 2018; Svaldi et al. 2021). Svaldi et al. (2021) employed a dual approach whereby FC data were first subjected to a principal component analysis (PCA)-based procedure aimed at improving participant identifiability. They then showed that the new FC matrices resulted in improved CPM performance to predict AD-relevant cognitive measures. Interestingly, Mantwill et al. (2022) recently showed that (at least in the young and healthy Human Connectome Project cohort) identifiability and behavior prediction are reliant upon distinct functional systems. Given this evidence it is unclear how a PCA-based improvement of FC aimed at identifiability impacts behavioral prediction. Another PCA-based approach by Shine et al. (2019), has been employed to separate the temporal BOLD signal into components, essentially weighting the each timepoint of a scan, examining for associations with task performance. Unlike the abovementioned methods, our application is simple in its selection of time points and statistical inference at RSN/block level. We hypothesized that particular time points may be more relevant to certain neuropsychological outcomes over others, thereby parsing them into bins based on co-fluctuation magnitude to estimate FC components (Cutts et al. 2022).

We conducted an exploratory analysis where a RSN block structure was not imposed on FC components. Treating the whole network as a single block, we looked for connected components that significantly correlated with neuropsychological domains using the network-based statistics correction strategy (Zalesky et al. 2010). As with the NCC strategy only, the cognitive function domain was correlated with FC, composed of a component that included the ventral attention network and its interactions with frontoparietal and default mode networks. Upper middle RSS bin components revealed significant components that seem to differ in their spatial distribution for each neuropsychological domain (Figure 5 shows primarily (1) ventral attention, frontoparietal and default mode relationships with cognitive, (2) visual, somatomotor, and dorsal attention interactions with other systems for executive, and (3) dorsal attention for language neuropsychological domains).

Our supplemental analyses where (1) FCc were generated from 10 bins of data and (2) a finer scale 300-node parcellation was used, showed similar findings to those reported in the main manuscript. For the latter analysis, scale is a particularly important factor as both statistical analysis frameworks (NCC and NBS) can be influenced by number of nodes in the network. For NCC, while in many cases the same blocks were identified, there were differences i.e., the results obtained for the FC and Cognitive domain interrelationships, where in the 200-node network visual and ventral attention system associations were identified, while in the 300-node network ventral attention-limbic interaction and limbic-default mode interaction blocks were significant. Among the 7 RSN systems, ventral attention and limbic systems show one of the higher increases in node count (35-40% increase), thus greatly increasing the number of edges.

Alternatively, the dorsal attention and frontoparietal systems showed the smallest difference in node count between the two scales (∼25% increase). This distinction in the extent the number of nodes increases within each system at finer scale is the most likely explanation for variance in the result between the two parcellations.

It is important to consider these findings within the constraints of the methodology and analyses employed. First, this is a cross-sectional investigation with the sample spanning the AD diagnostic continuum, aimed at investigating how FC components are altered in relation to disease-relevant neuropsychological domains. Future longitudinal follow-ups are necessary to assess whether this strategy reveals similar relationships in within-subject designs. Second, we chose a 200-region (and 300-region in supplement) functional cortical parcellation (Schaefer et al. 2018) stratified into seven canonical RSNs (Yeo et al. 2011). Whether the same or similar systems are implicated utilizing different parcellation and network stratifications can be a topic of future investigations, as node selection is often debated in network neuroscience and can be a source of variance in network data (Domhof et al. 2021). Additional inclusion of subcortical, cerebellar, and brainstem regions may shed light on relationships between neuropsychological function and interactions between cortical systems and the subcortex, however, this will require alternative statistical methods as it is not appropriate to treat the subcortex as a unitary block in a matrix. Finally, because of the frequency statistic-based testing of block-level relationships employed by NCC, we cannot, or rather should not, conduct follow-up tests to isolate the significant edges within blocks. Therefore, this method is limited in its interpretation as to whether or not there is a coupling between FC and behavior for a set of subsystems (here RSNs) only.

In summary, we hypothesized that a decomposition of FC into components derived from temporally discrete data points via an edge time series summary metric that indexes magnitude of co-fluctuation in a network will reveal brain-behavior relationships not observed with conventional full FC. Applied to a sample that spans the AD diagnostic spectrum, we show that discrete FC components are related to neuropsychological domain performance within and between specific RSN systems. This work can serve as a starting point for more targeted investigations of specific brain systems and how they relate to phenotypic changes as a consequence of AD and related dementias.

## Data Availability

Data access can be obtained by contacting the Indiana Alzheimer's Disease Center Data Core.

## Declarations

### Ethical Approval

Informed consent was obtained from all participants or their representatives, and all procedures were approved by the Indiana University Institutional Review board in accordance with the Belmont report.

### Author Contributions

**Evgeny J. Chumin**: Conceptualization, Methodology, Formal analysis, Software, Writing - original draft, Writing - review & editing. **Sarah A. Cutts**: Conceptualization, Methodology, Writing - review & editing. **Shannon L. Risacher**: Methodology, Resources, Data curation, Funding acquisition, Writing - review & editing. **Liana G. Apostolova**: Methodology, Funding acquisition, Writing - review & editing. **Martin R. Farlow**: Writing - review & editing. **Brenna C. McDonald**: Writing - review & editing. **Yu-Chien Wu**: Writing - review & editing, Funding acquisition. **Richard Betzel**: Conceptualization, Methodology, Writing - review & editing. **Andrew J. Saykin**: Supervision, Funding acquisition, Writing - review & editing. **Olaf Sporns**: Conceptualization, Methodology, Software, Supervision, Writing - review & editing.

### Data Availability

Data access can be obtained by contacting the Indiana Alzheimer’s Disease Center Data Core.

## Acknowledgements

The authors thank Eileen Tallman, Aaron Vosmeier, Rachael Deardorff, Bradley Glazier, Kala Hall, Lili Kyurkchiyska, Evan Finley, Yolanda Graham-Dotson, Steve Brown, and Sujuan Gao for their contributions to this study. We also thank the participants in the Indiana Memory and Aging Study (IMAS) and their family members, without whom this research would not be possible. The authors also acknowledge the Indiana University Pervasive Technology Institute (Stewart et al. 2017) for providing supercomputing and storage resources, supported in part by Lilly Endowment, Inc., through its support for the Indiana University Pervasive Technology Institute.

