## supplemental figures for "Edge Time Series Components of Functional Connectivity and Cognitive Function in Alzheimer’s Disease"

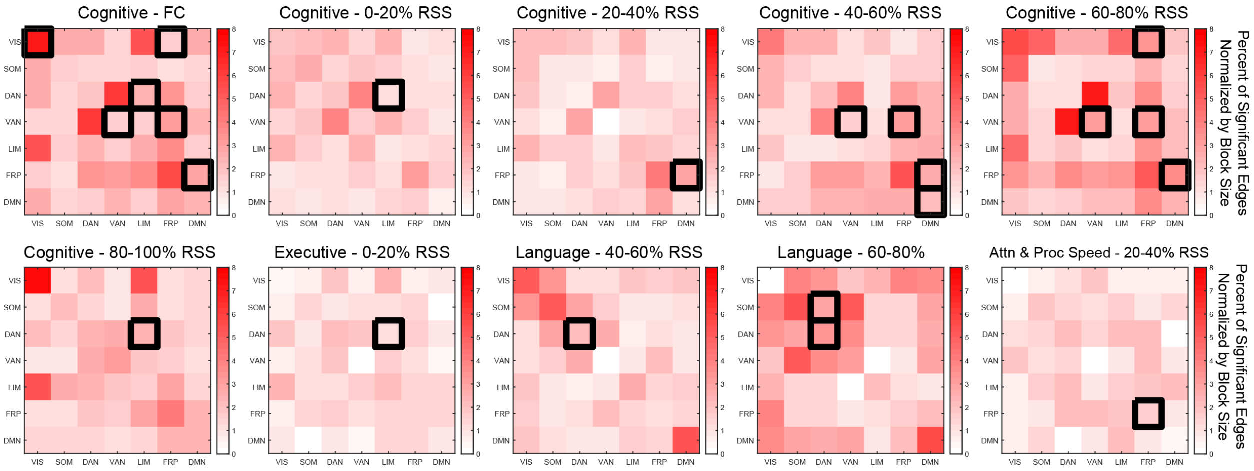


**Supplementary Figure 1. Significant blocks as assessed with the neuropsychological domain score scrambling null model.**

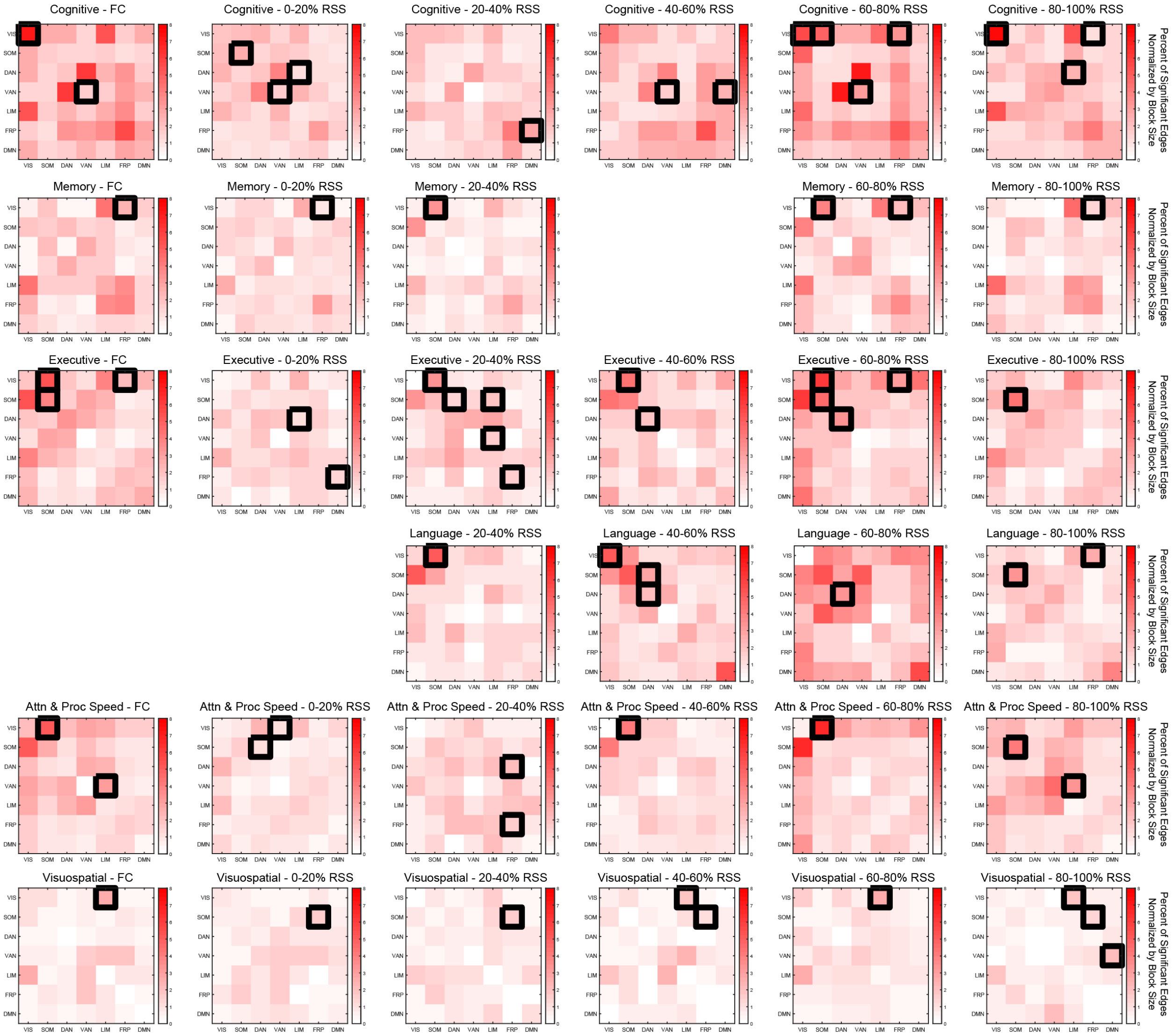


**Supplementary Figure 2. Significant blocks as assessed with the resting state network block structure scrambling null model.**

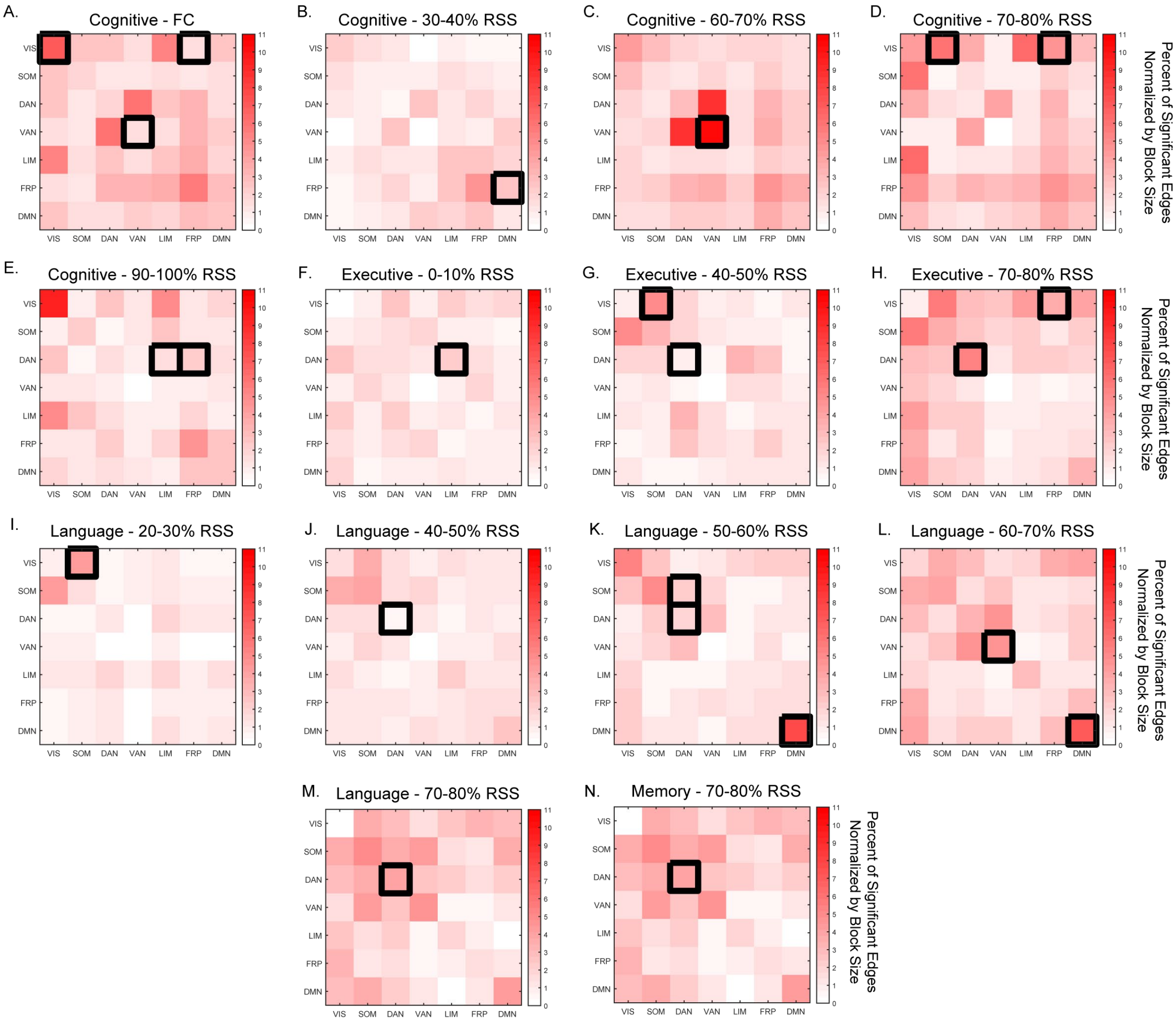


**Supplementary Figure 3. Significant blocks assessed from a decile split into ten FCcs. Only blocks that were significant in both null models are reported here.**

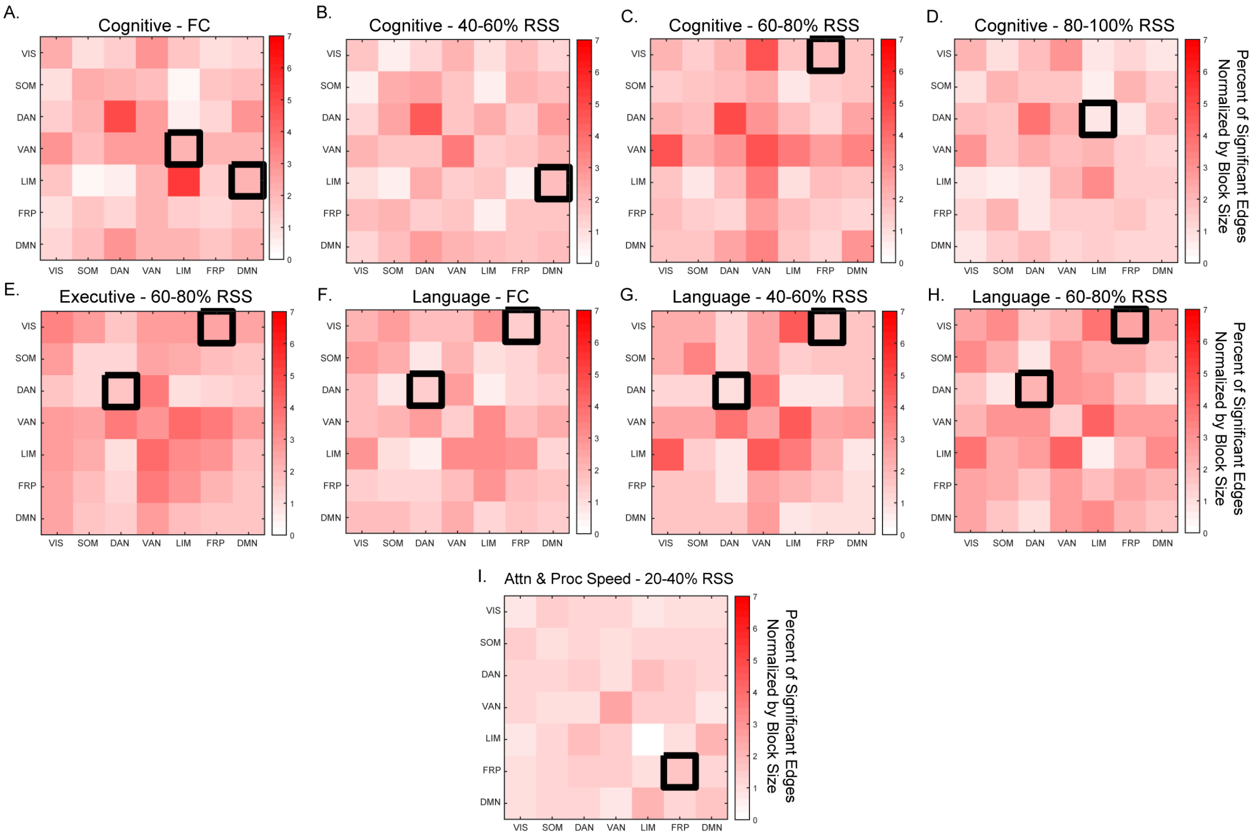


**Supplementary Figure 4. Significant blocks assessed form a Schaefer 300 node parcellation data split into 5 bin FCcs. Only blocks that were significant in both null models are reported here.**

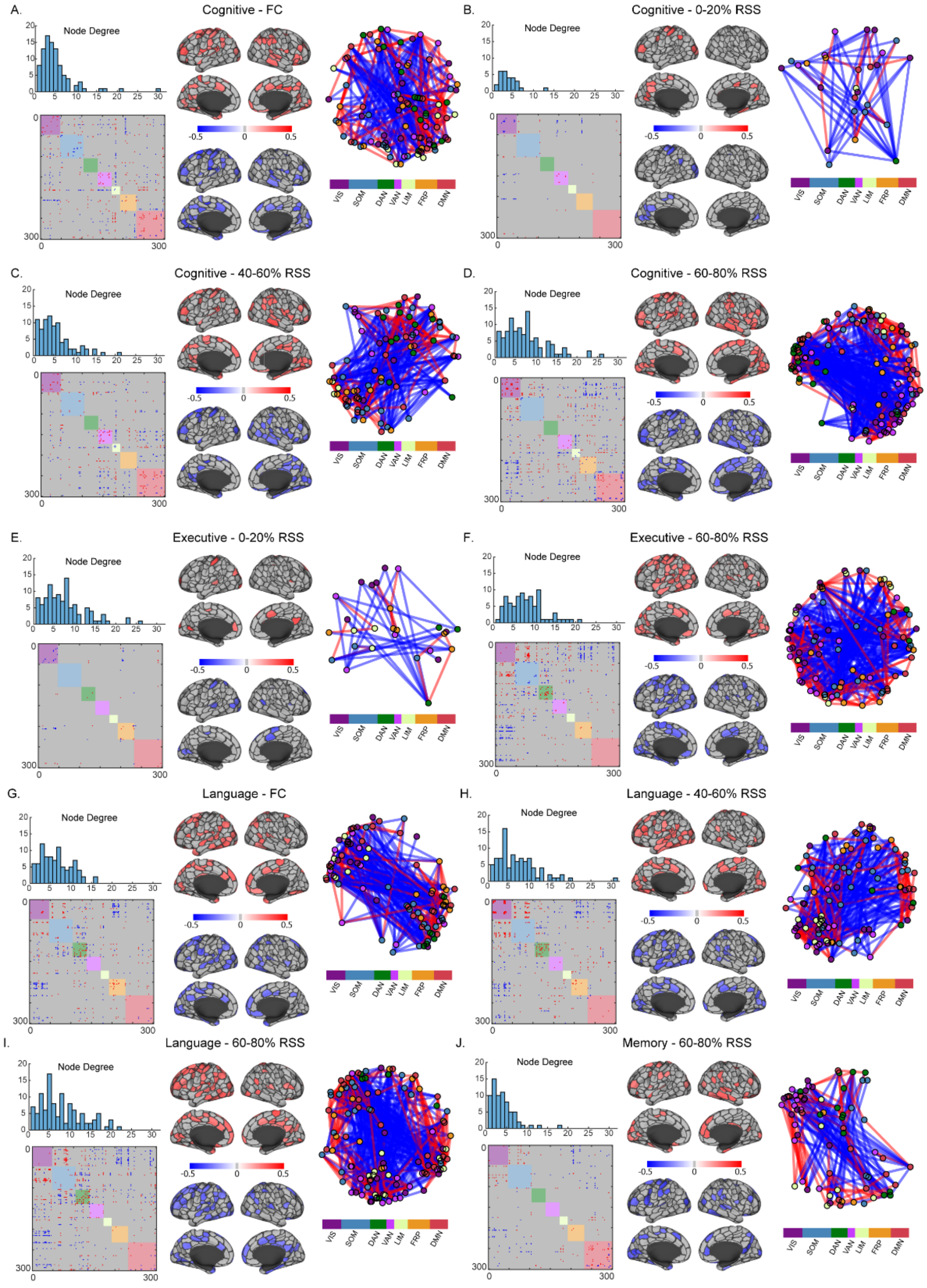


**Supplementary Figure 5. Block-fee Network-Based Statistics significant FC component-neuropsychological domain correlations from the Schaefer 300 node parcellated data and five RSS binned FC components.**
